# Socio-demographic Correlates of Prolonged Amenorrhea and Menopausal Transition among Nigerian Women Aged 30-49: Evidence from the 2024 Nigeria Demographic and Health Survey

**DOI:** 10.64898/2026.06.06.26355063

**Authors:** Olubusayo Bankole Ogunsemoyin, Aboluwaji Daniel Ayinmoro

## Abstract

**Introduction:** Menopause is a central marker of reproductive ageing, but national evidence on menstrual cessation among Nigerian women in the late reproductive ages remains limited. This study examined the prevalence and socio-demographic correlates of prolonged amenorrhea/possible menopausal transition among Nigerian women aged 30-49 years.

**Methods:** The study used the women’s individual recode file from the 2024 Nigeria Demographic and Health Survey. The analytic sample was restricted to women aged 30-49 years, excluding women who were currently pregnant, currently or postpartum amenorrheic, and those with invalid or special responses on time since last menstrual period. The final sample comprised 14,223 women. The outcome combined women whose last menstrual period occurred 12 or more months before the survey, and women reported as being in menopause. Weighted descriptive statistics, design-adjusted bivariate tests and survey-weighted binary logistic regression were used.

**Results:** The weighted prevalence of prolonged amenorrhea/possible menopausal transition was 7.6%. Prevalence rose from 1.2% among women aged 30-34 years to 23.6% among women aged 45-49 years. In the adjusted model, women aged 35-39 years (OR=1.64; p=0.030), 40-44 years (OR=6.20; p<0.001) and 45-49 years (OR=24.51; p<0.001) had higher odds than women aged 30-34 years. Primary education (OR=1.65; p=0.004), middle wealth status (OR=1.37; p=0.043) and poorest wealth status (OR=1.60; p=0.024) were associated with higher odds. Muslim affiliation (OR=0.72; p=0.024) and traditional contraceptive use (OR=0.24; p<0.001) were associated with lower odds.

**Conclusion:** Prolonged amenorrhea/possible menopausal transition among Nigerian women aged 30-49 is strongly age-patterned and socially differentiated. The findings support the need to make midlife menstrual health more visible within reproductive, family planning and primary healthcare services. Because the measure is based on survey-reported menstrual recency, it should not be interpreted as clinically confirmed natural menopause.

## Introduction

Menopause is a normal reproductive-ageing event, but its demographic and public health implications extend beyond the permanent cessation of menstruation. It marks the end of spontaneous reproductive capacity and is linked to broader endocrine, metabolic and later-life health changes [1-4]. The World Health Organisation defines natural menopause as occurring after 12 consecutive months without menstruation, with no other obvious physiological or pathological cause and no clinical intervention [2]. This distinction matters because amenorrhoea among women of reproductive age may also reflect pregnancy, lactation, postpartum amenorrhoea, contraceptive effects, illness, stress, hysterectomy or reporting error.

In population surveys, menopause is rarely measured with the clinical precision implied by this definition. The Stages of Reproductive Ageing Workshop and STRAW+10 frameworks distinguish late reproductive life, the menopausal transition, the final menstrual period and postmenopause, and emphasise that the final menstrual period can only be confirmed retrospectively after 12 months of amenorrhoea [3,4]. Cross-sectional surveys, including the Demographic and Health Surveys (DHS), instead rely on menstrual recency, pregnancy status, postpartum status and self-reported menopause or hysterectomy information. The DHS menopause indicator was designed primarily to measure biological non-exposure to pregnancy among women aged 30-49, and the DHS guide treats six months without a period, in the absence of pregnancy and postpartum amenorrhoea, as an indication of menopausal status in older reproductive-age women [5].

The present study adopts a more conservative approach. It uses the DHS variable for time since the last menstrual period (V226) but applies a 12-month threshold after excluding currently pregnant women and those with current or postpartum amenorrhoea. The outcome is therefore described as prolonged amenorrhoea/possible menopausal transition rather than confirmed menopause. This wording is deliberate. It preserves the demographic value of the NDHS while avoiding the clinical overstatement that would arise from equating a cross-sectional menstrual recency variable with natural menopause.

International evidence shows that the timing of reproductive ageing is socially patterned. A systematic review of studies across six continents indicates that age at natural menopause varies across populations and is associated with socio-economic position, education, occupation and lifestyle factors [6]. Multiethnic cohort evidence has also linked age at natural menopause with race/ethnicity, education, marital status, smoking, parity and oral contraceptive history [7]. Reproductive history is central to this discussion: early menarche and nulliparity have been associated with an increased risk of premature and early natural menopause in pooled international analyses [8]. These patterns matter because premature or early menopause has been associated with increased risk of coronary heart disease, cardiovascular mortality and all-cause mortality. At the same time, the menopausal transition is increasingly viewed as a window for early prevention of cardiovascular and metabolic risk [9-11].

Evidence from low- and middle-income countries suggests that premature and early menopause are not marginal issues in poorer settings [12]. However, the evidence base in sub-Saharan Africa remains uneven, and Nigeria-specific studies remain largely subnational. Earlier Nigerian studies have reported menopausal ages ranging from the late forties to the early fifties, with variation by study site, parity, education, body mass index, occupation, and reproductive history [13-16]. These studies are useful but cannot replace a nationally representative analysis of menstrual recency among women aged 30-49 years.

Nigeria provides an important setting for this analysis because women aged 30-49 are at different stages of the reproductive life cycle. Some remain actively exposed to pregnancy, some are approaching the end of their reproductive lives, and others may already have entered the menopausal transition. These stages unfold within a context of high fertility heterogeneity, regional inequalities, uneven access to reproductive and primary healthcare, socio-economic stratification and cultural diversity [1]. Understanding the distribution and correlates of prolonged amenorrhoea in this age group has implications for fertility measurement, family planning counselling, menstrual health, chronic disease prevention and the visibility of midlife women in health policy.

This study, therefore, examined prolonged amenorrhoea/possible menopausal transition among Nigerian women aged 30-49 years using the 2024 NDHS. Specifically, it estimated the weighted prevalence of the outcome and assessed its socio-demographic and reproductive correlates, excluding pregnancy- and postpartum-related explanations available in the DHS file.

### Theoretical framework

The analysis is guided by an integrated framework that combines life-course epidemiology, the social determinants of health perspective, fundamental cause theory and reproductive-ageing staging. Life-course epidemiology argues that adult health reflects biological, behavioural, and social exposures accumulated from early life through later adulthood [17,18]. Applied to menstrual cessation, this perspective suggests that accumulated nutrition, infections, reproductive histories, physical workload, chronic stress and access to healthcare may shape reproductive ageing.

The social determinants of health perspective locates these exposures within unequal distributions of power, resources and opportunities [19,20]. Fundamental cause theory further argues that social conditions influence health because they embody flexible resources such as money, knowledge, prestige, power and beneficial social connections [21,22]. In this study, education, household wealth, residence, ethnicity, religion and work status are therefore interpreted as markers of social location rather than as direct biological causes. The STRAW/STRAW+10 framework provides the reproductive-ageing logic by treating menstrual cycle change and amenorrhoea as proximate markers of transition, while reserving clinical menopause for retrospectively confirmed 12-month amenorrhoea [3,4].

The article’s conceptual position is therefore straightforward: socio-structural location shapes reproductive life-course exposures; reproductive history and contraceptive context influence menstrual patterns; and V226 captures the survey expression of menstrual recency. The outcome is interpreted as prolonged amenorrhoea/possible menopausal transition, not as clinically verified natural menopause.

## Materials and methods

### Study design and data source

This study was a cross-sectional secondary analysis of the women’s individual recode file from the 2024 Nigeria Demographic and Health Survey (NDHS). The 2024 NDHS is a nationally representative household survey implemented by the Federal Ministry of Health and Social Welfare of Nigeria, the National Population Commission and ICF. It used a stratified multistage cluster sampling design. It collected information on fertility, reproductive history, contraceptive use, marital and union status, menstrual recency, employment and background characteristics among women of reproductive age [1]. The dataset was accessed for research purposes on 4 May 2026, following approval via the DHS Program data access procedure.

### Study population and sample construction

The target population comprised Nigerian women aged 30-49 years who were interviewed and had valid information on menstrual recency, after excluding pregnancy- and postpartum-related explanations for amenorrhoea. The working NDHS file contained 17,967 women aged 30-49 years. After these exclusions, the analysis excluded 1,195 currently pregnant women, 2,423 currently or postpartum amenorrhoeic women, and 126 women with invalid or special V226 responses. The final analytic sample comprised 14,223 women.

**Table 1.**
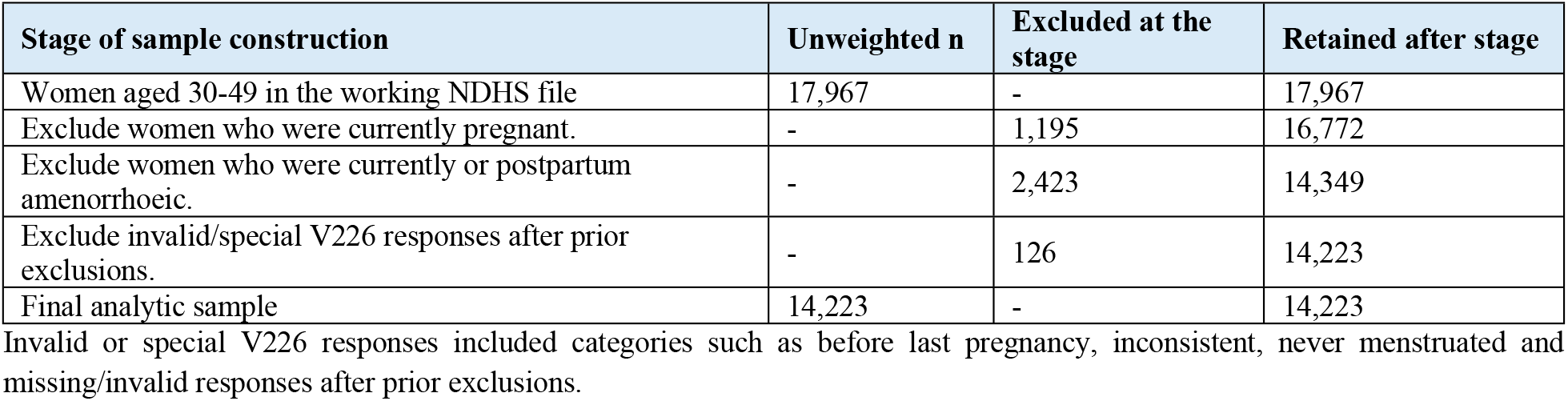
Sample construction for the V226-based analytic sample.

### Outcome variable

The outcome was derived from V226, the DHS variable measuring time since the last menstrual period. Women were coded 0 if their last menstrual period occurred 0-11 months before the survey. Women were coded 1 if their last menstrual period occurred 12 or more months before the survey, or if they were reported as being in menopause. This definition follows the 12-month criterion used in clinical and reproductive-ageing frameworks, while recognising that the NDHS does not clinically confirm natural menopause through biomarkers or longitudinal observation [2-4].

### Explanatory variables

Explanatory variables were selected based on theoretical relevance, prior literature and availability in the working file. Socio-demographic variables included age group, place of residence, highest level of education, religion, ethnicity, household wealth quintile and current work status. Reproductive variables included children ever born, age at first birth, marital/union status and current contraceptive method. Current contraceptive methods were grouped into not using, modern methods and traditional methods. These variables captured social position, reproductive life-course exposure and proximate menstrual context.

### Statistical analysis

Analyses accounted for the NDHS complex survey design. The women’s individual sampling weight (V005) was divided by 1,000,000 before analysis. Weighted percentages were used for descriptive estimates, while unweighted counts were retained to indicate the observed analytic sample size. Bivariate associations between explanatory variables and prolonged amenorrhoea/possible menopausal transition were assessed using design-adjusted tests. Survey-weighted binary logistic regression was then used to estimate adjusted odds ratios and 95% confidence intervals. Statistical significance was assessed at the 5% level. Exact p-values are reported where available.

### Ethics statement

This study used de-identified secondary data from the DHS Program. The 2024 NDHS protocol received ethical approval from relevant national and institutional review bodies, and informed consent was obtained from respondents prior to data collection [1]. No new primary data were collected for this analysis, and the authors did not use respondent-identifying information. Access to the dataset was obtained in accordance with the DHS Program’s data-use requirements.

## Results

### Socio-demographic and reproductive characteristics of respondents

Table 2 presents the weighted socio-demographic and reproductive characteristics of the analytic sample. The sample was fairly evenly distributed across the age groups 30-34, 35-39 and 40-44, while women aged 45-49 accounted for a smaller share. Slightly more than half of the women lived in urban areas. About one-third had no formal education, and another one-third had secondary education. The sample was almost evenly split between Muslim and Christian women, with very few women reporting traditional or other religious affiliations.

**Table 2.** Weighted socio-demographic and reproductive characteristics of women aged 30-49.

| Variable | Category | Unweighted (n) | Weighted (%) |
| --- | --- | --- | --- |
| Age group | 30-34 | 3,672 | 26.3 |
|  | 35-39 | 3,837 | 26.9 |
|  | 40-44 | 3,632 | 25.7 |
|  | 45-49 | 3,082 | 21.1 |
| Place of residence | Urban | 7,502 | 53.0 |
|  | Rural | 6,721 | 47.0 |
| Highest level of education | No education | 4,403 | 34.5 |
|  | Primary | 2,085 | 13.9 |
|  | Secondary | 4,995 | 33.1 |
|  | Higher | 2,740 | 18.6 |
| Religion | Islam | 5,917 | 48.7 |
|  | Christianity | 8,215 | 50.8 |
|  | Traditional/Other | 91 | 0.5 |
| Ethnicity | Hausa/Fulani | 3,488 | 30.2 |
|  | Yoruba | 2,286 | 17.4 |
|  | Igbo | 2,844 | 15.2 |
|  | Northern minority | 3,351 | 23.9 |
|  | Southern minority | 2,007 | 11.8 |
|  | Unclassified | 247 | 1.6 |
| Household wealth quintile | Poorest | 2,075 | 14.4 |
|  | Poorer | 2,057 | 16.1 |
|  | Middle | 2,753 | 18.8 |
|  | Richer | 3,307 | 21.9 |
|  | Richest | 4,031 | 28.8 |
| Children ever born | 0 | 965 | 6.3 |
|  | 1-4 | 6,988 | 48.2 |
|  | 5+ | 6,270 | 45.5 |
| Age at first birth | No birth | 965 | 6.3 |
|  | <18 | 3,038 | 22.5 |
|  | 18-24 | 6,565 | 46.3 |
|  | 25+ | 3,655 | 24.9 |
| Marital/union status | Never in union | 779 | 4.9 |
|  | Currently in a union/living with a man | 11,919 | 85.0 |
|  | Formerly in a union/living with a man | 1,525 | 10.1 |
| Current contraceptive method | Not using | 10,261 | 72.9 |
|  | Modern method | 2,851 | 19.8 |
|  | Traditional method | 1,111 | 7.3 |
| Current work status | Currently working | 10,997 | 76.2 |
|  | Not currently working | 3,226 | 23.8 |

Percentages are weighted using V005/1,000,000; counts are unweighted. Source: 2024 NDHS women's individual recode file.

The reproductive profile reflected substantial marital and childbearing exposure. Most women were currently in a union or living with a man, nearly half had one to four children, and a similar proportion had five or more children. Age at first birth was concentrated between 18 and 24 years, although early first birth before age 18 remained common. Nearly three-quarters of women were not using contraception, about one-fifth used a modern method, and the use of traditional methods was uncommon. Most women were currently working.

### Prevalence of prolonged amenorrhea/possible menopausal transition

Table 3 shows the weighted prevalence of prolonged amenorrhoea/possible menopausal transition by age group. Overall, 92.4% of women reported that their last menstrual period occurred within 0-11 months before the survey, 5.5% reported 12 or more months since their last period, and 2.0% were reported as being in menopause. When the latter two categories were combined, the weighted prevalence of prolonged amenorrhoea/possible menopausal transition was 7.6%.

**Table 3.** Weighted prevalence of prolonged amenorrhea/possible menopausal transition by age group.

| Age group | Unweighted (n) | 0-11 months since last period (%) | 12+ months since last period (%) | Reported in menopause (%) | Combined outcome (%) |
| --- | --- | --- | --- | --- | --- |
| Total | 14,223 | 92.4 | 5.5 | 2.0 | 7.6 |
| 30-34 | 3,672 | 98.8 | 1.2 | 0.0 | 1.2 |
| 35-39 | 3,837 | 98.1 | 1.8 | 0.1 | 1.9 |
| 40-44 | 3,632 | 93.1 | 5.5 | 1.5 | 6.9 |
| 45-49 | 3,082 | 76.4 | 15.9 | 7.7 | 23.6 |

The combined outcome includes women with 12 or more months since their last menstrual period, and women reported as being in menopause. Percentages are weighted; counts are unweighted.

The prevalence varied sharply by age. Prolonged amenorrhea/possible menopausal transition was uncommon before age 40, increased among women aged 40-44, and rose most strongly among women aged 45-49. This age pattern is consistent with reproductive ageing. However, the small proportions among younger women require cautious interpretation because they may include early transition, non-menopausal amenorrhea, residual reporting error or other reproductive health conditions.

### Bivariate analysis

Table 4 presents bivariate associations between explanatory variables and prolonged amenorrhoea/possible menopausal transition.

**Table 4.**
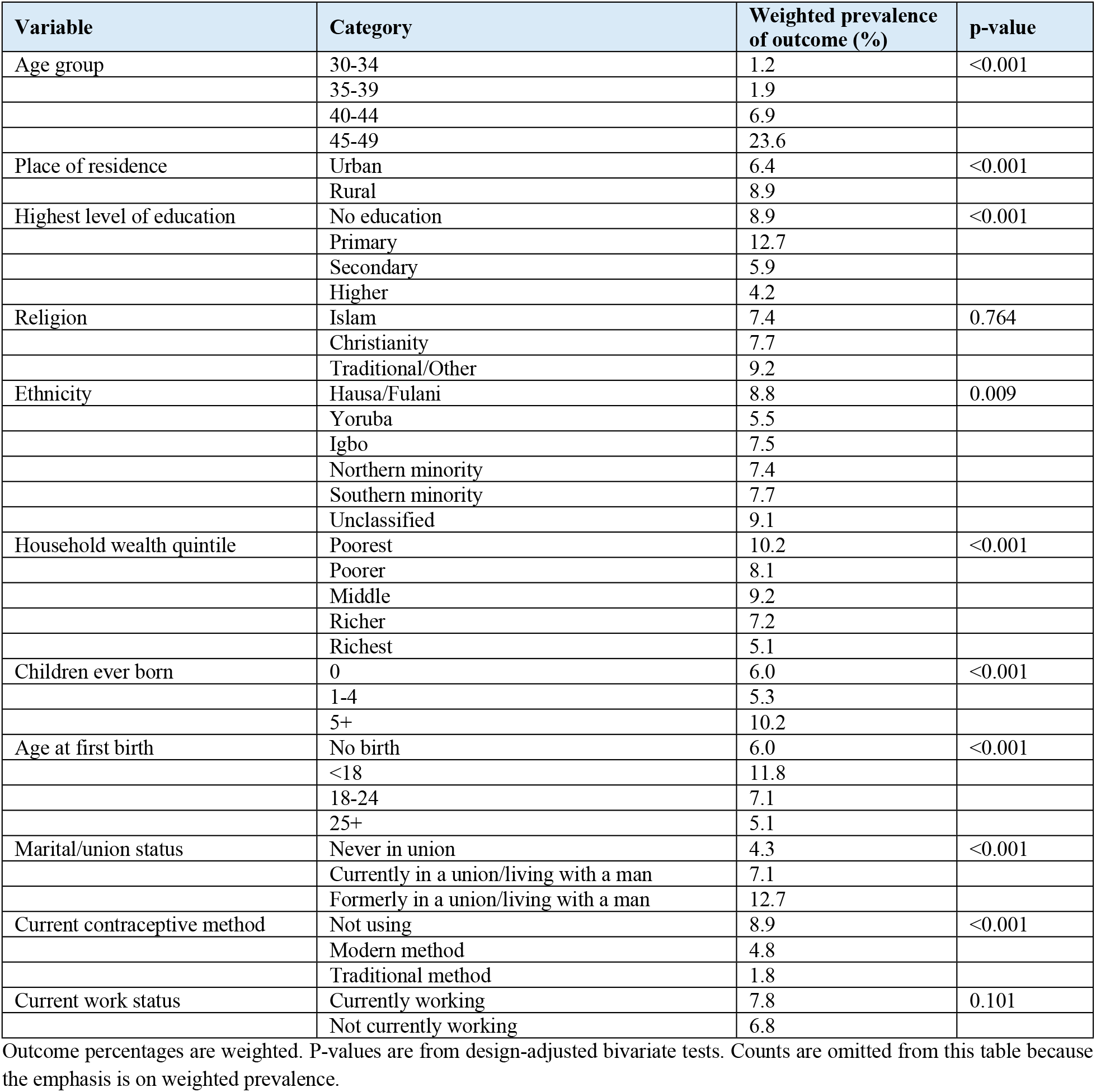
Bivariate association between selected characteristics and prolonged amenorrhea/possible menopausal transition.

The outcome varied significantly by age group, place of residence, education, ethnicity, household wealth, children ever born, age at first birth, marital/union status and current contraceptive method. Religion and current work status were not statistically significant at the bivariate level. The prevalence increased steeply with age and was higher among rural than among urban women. By education, the outcome was most common among women with primary education and least common among those with higher education. Ethnic variation was observed, with the highest prevalence among Hausa/Fulani women and the lowest among Yoruba women. A wealth gradient was also evident: women in the poorest households had the highest prevalence, while those in the richest had the lowest.

Reproductive characteristics showed clear variation. Women with five or more children had a higher prevalence than those with fewer children or no children. Women whose first birth occurred before age 18 had the highest prevalence, while those whose first birth occurred at age 25 or later had the lowest. Formerly married or cohabiting women had a higher prevalence than those currently in union and those never in union. By contraceptive status, prevalence was highest among non-users and lower among users of modern or traditional methods.

### Adjusted correlates of prolonged amenorrhea/possible menopausal transition

Table 5 presents the results of the adjusted logistic regression. Age was the dominant predictor. Compared with women aged 30-34, those aged 35-39 had significantly higher odds of prolonged amenorrhoea/possible menopausal transition. The odds increased substantially among women aged 40-44 and were highest among those aged 45-49.

**Table 5.**
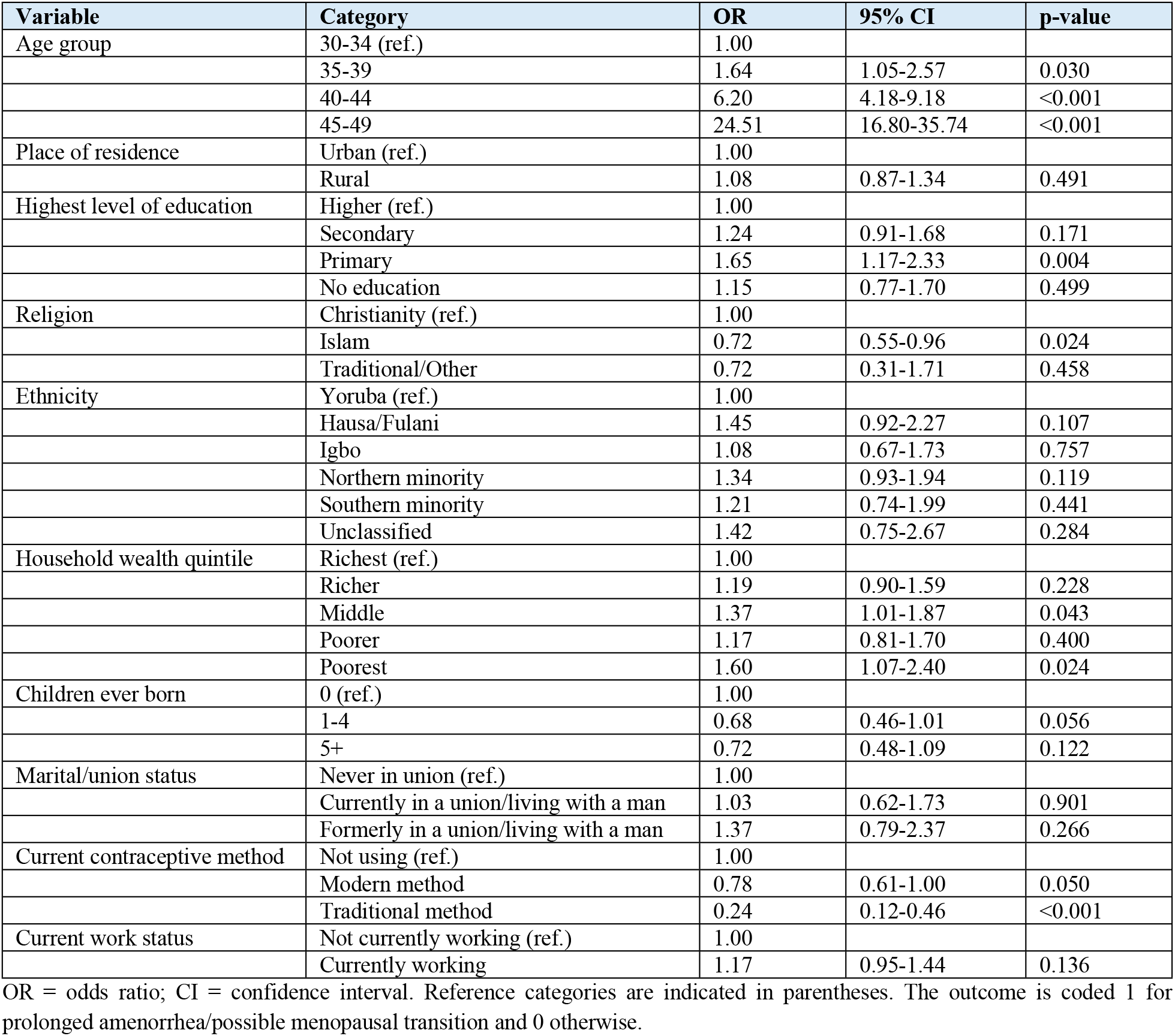
Survey-weighted logistic regression of prolonged amenorrhea/possible menopausal transition among women aged 30-49.

| Variable | Category | OR | 95% CI | p-value |
| --- | --- | --- | --- | --- |
| Age group | 30-34 (ref.) | 1.00 |  |  |
|  | 35-39 | 1.64 | 1.05-2.57 | 0.030 |
|  | 40-44 | 6.20 | 4.18-9.18 | <0.001 |
|  | 45-49 | 24.51 | 16.80-35.74 | <0.001 |
| Place of residence | Urban (ref.) | 1.00 |  |  |
|  | Rural | 1.08 | 0.87-1.34 | 0.491 |
| Highest level of education | Higher (ref.) | 1.00 |  |  |
|  | Secondary | 1.24 | 0.91-1.68 | 0.171 |
|  | Primary | 1.65 | 1.17-2.33 | 0.004 |
|  | No education | 1.15 | 0.77-1.70 | 0.499 |
| Religion | Christianity (ref.) | 1.00 |  |  |
|  | Islam | 0.72 | 0.55-0.96 | 0.024 |
|  | Traditional/Other | 0.72 | 0.31-1.71 | 0.458 |
| Ethnicity | Yoruba (ref.) | 1.00 |  |  |
|  | Hausa/Fulani | 1.45 | 0.92-2.27 | 0.107 |
|  | Igbo | 1.08 | 0.67-1.73 | 0.757 |
|  | Northern minority | 1.34 | 0.93-1.94 | 0.119 |
|  | Southern minority | 1.21 | 0.74-1.99 | 0.441 |
|  | Unclassified | 1.42 | 0.75-2.67 | 0.284 |
| Household wealth quintile | Richest (ref.) | 1.00 |  |  |
|  | Richer | 1.19 | 0.90-1.59 | 0.228 |
|  | Middle | 1.37 | 1.01-1.87 | 0.043 |
|  | Poorer | 1.17 | 0.81-1.70 | 0.400 |
|  | Poorest | 1.60 | 1.07-2.40 | 0.024 |
| Children ever born | 0 (ref.) | 1.00 |  |  |
|  | 1-4 | 0.68 | 0.46-1.01 | 0.056 |
|  | 5+ | 0.72 | 0.48-1.09 | 0.122 |
| Marital/union status | Never in union (ref.) | 1.00 |  |  |
|  | Currently in a union/living with a man | 1.03 | 0.62-1.73 | 0.901 |
|  | Formerly in a union/living with a man | 1.37 | 0.79-2.37 | 0.266 |
| Current contraceptive method | Not using (ref.) | 1.00 |  |  |
|  | Modern method | 0.78 | 0.61-1.00 | 0.050 |
|  | Traditional method | 0.24 | 0.12-0.46 | <0.001 |
| Current work status | Not currently working (ref.) | 1.00 |  |  |
|  | Currently working | 1.17 | 0.95-1.44 | 0.136 |

OR = odds ratio; CI = confidence interval. Reference categories are indicated in parentheses. The outcome is coded 1 for prolonged amenorrhea/possible menopausal transition and 0 otherwise.

Place of residence was not significantly associated with the outcome after adjustment. Education showed a non-linear pattern. Women with primary education had significantly higher odds than those with higher education. In contrast, women with secondary education and those with no education did not differ significantly from the higher education category. Muslim women had significantly lower odds than Christian women, while women reporting traditional or other religious affiliations did not differ significantly from Christian women. Ethnicity was not independently associated with the outcome after adjustment.

Household wealth remained partially associated with the outcome. Women in the middle and poorest wealth quintiles had significantly higher odds than those in the richest quintile.

Children ever born and marital/union status were not independently associated with prolonged amenorrhoea/possible menopausal transition. The current contraceptive method was associated with the outcome: traditional method users had substantially lower odds than non-users, whereas the association for modern method use was borderline. The current work status was not statistically significant.

## Discussion

This study examined prolonged amenorrhoea/possible menopausal transition among Nigerian women aged 30-49 years using the 2024 NDHS. The findings show that the outcome is strongly age-patterned, socially differentiated, and sensitive to reproductive and contraceptive context. Overall prevalence was 7.6%, but it rose sharply across age groups, reaching nearly one-quarter among women aged 45-49. This age gradient supports the reproductive-ageing interpretation of the outcome while reinforcing the need for caution among younger women, where prolonged amenorrhoea may reflect causes other than menopausal transition.

The strong age gradient is consistent with STRAW+10, which places menstrual variability and amenorrhoea within the sequence of reproductive ageing [3]. It also aligns with Nigerian studies showing that menopause commonly occurs in the late forties or early fifties [13-16]. However, this study differs from earlier Nigerian work because it does not estimate age at natural menopause among women already known to be menopausal. Instead, it describes a nationally representative distribution of prolonged menstrual absence among women aged 30-49, after excluding pregnancy- and postpartum-related explanations available in the DHS file. This makes the study useful for demographic measurement, but it also limits clinical interpretation.

The bivariate results showed rural, educational, wealth, ethnic and reproductive differentials. Several of these associations weakened after adjustment, as expected in a setting where residence, ethnicity, religion, education, parity and wealth are strongly interrelated. The disappearance of residence and ethnicity effects in the adjusted model suggests that their bivariate associations may reflect compositional differences in age, socio-economic position and reproductive history rather than independent effects. This interpretation is consistent with social determinants and fundamental cause perspectives, which treat social categories as markers of structured exposure rather than isolated causal agents [19-22].

The educational result is important but should not be overinterpreted. Women with primary education had higher adjusted odds than those with higher education, whereas women with no education did not differ significantly from those with higher education. This non-linear pattern may reflect cohort composition, reporting differences, residual confounding or heterogeneity within the no-education category. It should not be read as evidence that lack of schooling protects women from prolonged amenorrhoea. Rather, it suggests that educational gradients in menstrual cessation are not necessarily monotonic once age, wealth, religion, ethnicity and reproductive factors are considered.

The wealth pattern is more consistent with a life-course interpretation. Women in the poorest and middle wealth quintiles had higher odds than those in the richest quintile. Lower household wealth may reflect accumulated disadvantage, poorer nutrition, reduced access to preventive care, and greater exposure to physically demanding conditions. These pathways align with life-course evidence linking social and environmental conditions across the life course to the timing of menopause [17,23,24]. However, causality cannot be inferred from the cross-sectional design, and household wealth measured at the time of the survey may not fully capture earlier-life deprivation.

The lower adjusted odds among Muslim women compared with Christian women should also be interpreted cautiously. In Nigeria, religion is closely tied to region, ethnicity, marriage systems, fertility histories and healthcare access. The finding may therefore reflect a conditional association after adjustment rather than a direct religious effect. Similarly, the absence of an independent ethnicity effect after adjustment should not be taken as evidence that cultural context is irrelevant. Rather, broad ethnic categories may be too coarse to capture local meanings, health behaviours, and reporting practices related to menstrual cessation.

The contraceptive findings warrant caution. Traditional method users had substantially lower odds of prolonged amenorrhoea/possible menopausal transition, and modern method users had marginally lower odds than non-users. Traditional methods do not biologically prevent menopause; therefore, the inverse association is more plausibly explained by selection into method use, continued perceived fertility, age composition, sexual exposure, menstrual regularity, or classification effects. The broad modern-method category also combines methods with different effects on bleeding. A method-specific analysis would be needed to distinguish hormonal methods that may induce amenorrhoea from non-hormonal modern methods.

The study contributes to the literature in two ways. First, it provides nationally representative evidence from Nigeria on prolonged menstrual absence among women aged 30-49, rather than relying on subnational or facility-based samples. Second, it reframes midlife menstrual cessation as both a reproductive-ageing and a social-equity issue. Menopause and the menopausal transition are often underemphasised in reproductive health programmes, which tend to prioritise fertility regulation, pregnancy, childbirth and adolescent health. The results suggest that Nigerian women in the late reproductive years require more explicit counselling and services on menstrual changes, contraceptive interpretation, chronic disease screening and referral for abnormal amenorrhoea.

## Strengths and limitations

A major strength of this study is the use of a nationally representative dataset with survey weights and design variables, which supports population-level inference for Nigerian women aged 30-49. The study also uses V226 directly, anchoring the outcome to menstrual recency. Excluding currently pregnant women, currently or postpartum amenorrhoeic women, and invalid or special V226 responses reduces the risk of misclassifying pregnancy- and postpartum-related menstrual absence as menopausal transition.

The study also has limitations. V226 is self-reported and does not provide clinical confirmation of natural menopause. The cross-sectional design prevents prospective observation of the final menstrual period and precludes causal inference. Relevant determinants, such as smoking, body mass index, nutrition, chronic illness, hysterectomy, oophorectomy, family history and detailed method-specific hormonal exposure, were not included in the presented model. Residual misclassification is therefore possible, particularly among younger women classified as having prolonged amenorrhoea. Finally, broad categories for ethnicity, religion and contraceptive method may obscure important within-group variation.

## Implications for policy and research

The findings indicate that midlife menstrual health should be more visible across reproductive, family planning and primary healthcare services in Nigeria. Primary healthcare workers, family planning providers and community health educators should be able to counsel women on expected menstrual changes, signs that require referral, and the differences between reproductive ageing, pregnancy, contraceptive-related bleeding changes and abnormal amenorrhoea. Counselling is particularly important for women aged 40-49, but younger women reporting prolonged menstrual absence should also be assessed for pregnancy, postnatal status, contraceptive effects and possible pathological causes.

Future research should examine method-specific contraceptive effects, incorporate body mass index and smoking status where available, and model regional variation. Longitudinal or clinical studies are needed to distinguish natural menopause from other causes of amenorrhoea. Qualitative work would also be useful for understanding how Nigerian women interpret, report and manage prolonged menstrual absence across diverse social and cultural settings.

## Conclusion

This study provides nationally representative evidence on prolonged amenorrhoea/possible menopausal transition among Nigerian women aged 30-49 years, using the 2024 NDHS. The prevalence was 7.6% overall and rose sharply with age, reaching nearly one-quarter among women aged 45-49. After adjustment, age remained the dominant predictor, while primary education, household wealth, Muslim affiliation and current contraceptive method also showed independent associations. The findings reinforce the need to treat midlife menstrual cessation as a demographic, reproductive health and social equity issue. Because the outcome is based on survey-reported menstrual recency, it should be interpreted as prolonged amenorrhoea/possible menopausal transition rather than clinically confirmed natural menopause.

## Data Availability

The study used publicly available, anonymised secondary data from the 2024 Nigeria Demographic and Health Survey. The dataset is available from The DHS Program upon registration and approval for legitimate research purposes. The specific dataset used was the Nigeria: Standard DHS, 2024 Women’s Recode file. Interested researchers can access the dataset through The DHS Program data portal at: https://dhsprogram.com/data/dataset/Nigeria_Standard-DHS_2024.cfm. The authors did not have any special access privileges that others would not have. The analytic dataset was derived from the publicly available DHS microdata using the eligibility restrictions and variable recoding described in the Methods section.

https://dhsprogram.com/data/dataset/Nigeria_Standard-DHS_2024.cfm?flag=0

## Notes

### Competing Interest Statement

The authors have declared no competing interest.

### Funding Statement

The author(s) received no specific funding for this work.

### Author Declarations

Ethical approval for the 2024 Nigeria Demographic and Health Survey was obtained by the National Health Research Ethics Committee of Nigeria and the ICF Institutional Review Board. The survey was conducted in accordance with standard DHS ethical procedures, and informed consent was obtained from participants before interview. The present study involved secondary analysis of anonymised, publicly available DHS data accessed with permission from The DHS Program. No additional primary data were collected, and the authors did not have access to information that could directly identify individual participants.

